# Inter-scanner Aβ-amyloid PET harmonization using barrel phantom spatial resolution matching

**DOI:** 10.1101/2023.07.16.23292733

**Authors:** Gihan P. Ruwanpathirana, Robert C. Williams, Colin L. Masters, Christopher C. Rowe, Leigh A. Johnston, Catherine E. Davey

## Abstract

The standardized uptake value ratio (SUVR) is used to measure Aβ uptake in PET images of the brain. Variations in PET scanner technologies and image reconstruction techniques can lead to variability in images acquired from different scanners. This poses a challenge for Aβ-PET studies conducted across multiple centers. The aim of harmonization is to achieve consistent Aβ-PET measurements across scanners. In this study, the procedure of matching the spatial resolution of a barrel phantom measured in each PET scanner is proposed as a method of Aβ-PET harmonization, validated using subject data.

**Methods:** Three different PET scanners were used: the Siemens Biograph Vision 600, Siemens Biograph mCT, and Philips Gemini TF64. A total of five, eight, and five subjects were each scanned twice with [^18^F]-NAV4694 across Vision-mCT, mCT-Philips, and Vision-Philips scanner pairs. The Vision and mCT scans were reconstructed using various iterations, subsets, and post-reconstruction Gaussian smoothing, while one reconstruction configuration was used for the Philips scans. The full-width at half-maximum (FWHM) of each reconstruction configuration was calculated using [^18^F]-filled barrel phantom scans with the SNMMI phantom analysis toolkit. Regional SUVRs were calculated from 72 brain regions using the AAL3 atlas for each subject and reconstruction configuration. Statistical similarity between SUVRs was assessed using paired (within subject) t-tests for each pair of reconstructions across scanners; the higher the p-value, the greater the similarity between the SUVRs.

**Results:** *Vision-mCT harmonization:* maximal statistical similarity (i.e., *p*-value) between regional SUVRs was achieved using a 4.10 mm FWHM Vision reconstruction with a 4.30 mm FWHM mCT reconstruction.

*Philips-mCT harmonization:* maximal statistical similarity between regional SUVRs was achieved using an 8.2 mm FWHM Philips reconstruction with a 9.35 mm FWHM mCT reconstruction.

*Philips-Vision harmonization:* a 9.1 mm FWHM Vision reconstruction had maximum statistical similarity with regional SUVRs from an 8.2mm FWHM Philips reconstruction. Reconstruction pairs that maximized statistical similarity, and supported a null hypothesis of being drawn from the same distribution, were selected as harmonised for each scanner pair.

**Conclusion:** Using data obtained from three sets of participants, each scanned on a different pair of PET scanners, using reconstruction configurations with matched barrel phantom spatial resolutions, we have demonstrated that Aβ-PET quantitation can be harmonised across scanners, producing SUVR values statistically likely to be drawn from the same distribution. This finding is encouraging for the use of different PET scanners in multi-centre trials, or updates during longitudinal studies.

## INTRODUCTION

The imaging of Amyloid beta (Aβ) protein plaques in the brain using positron emission tomography (PET) is of great importance in the study of Alzheimer’s disease, with the development of Aβ-specific radiotracers including [^18^F]-florbetaben, [^18^F]-florbetapir, [^18^F]-NAV4694 and [^18^F]-flutemetamol plaques enabling better monitoring of AD patients (*1*–*4*). While the extent of Aβ-PET accumulation in the brain is commonly measured using a standardized uptake value ratio (SUVR) (*2*), these values are known to be dependent on the choice of Aβ-PET radiotracer and scanner hardware (*5,6*). The spatial resolution of PET data is governed by physical factors including positron range of the radiotracer, photon scattering and hardware-specific limitations, and additionally the choice of reconstruction algorithm and associated parameter settings (*7,8*). It has been well established that PET quantitation is impacted by the partial volume effect (PVE), in which the intensity of a voxel is determined by both the dominant tissue within the voxel and the surrounding tissue that falls within the voxel. Therefore, choice of reconstruction parameters governing spatial resolution, and hence determining the PVE extent, influences Aβ quantitation (*6,9,10*). Furthermore, we have established that spatial resolution differentially impacts Aβ-PET quantitation in the Aβ- and Aβ+ cohorts (*7*). These scanner-dependent effects, in conjunction with the rapid advancement of PET technologies leading to significant performance variation across scanners, represent a problem for multi-site Aβ-PET studies; the same patient imaged on different PET scanners can exhibit significant image and Aβ-PET metric variability, even when attempts are made to match reconstruction parameters across scanning sites.

Problems associated with inter-scanner variability have long been recognised by the oncology community. The European Association of Nuclear Medicine (EANM) launched the EANM Research Ltd. (EARL) initiative to standardise images and promote multicentre studies (*12,13*). The EARL initiative employed the recovery coefficient, defined as the ratio of observed to true activity in PET, to compare PET reconstructions. They defined upper and lower limits for recovery coefficients using the National Electrical Manufacturers Association’s (NEMA) NU 2 body phantom imaging, and participating scanning sites choose a PET reconstruction configuration such that the recovery coefficients of their phantom images fall within the limits. EARL phantom criteria are not necessarily suitable for brain PET, as the NEMA NU 2 phantom contains multiple spheres with high radioactivity values within uniform background radioactivity. Although the NEMA NU 2 setup is suitable to simulate tumour uptake, it does not simulate brain uptake, which is characteristically different with a more dispersed uptake across larger grey matter and white matter compartments (*14*).

In the neuroimaging domain, Alzheimer’s Disease Neuroimaging Initiative (ADNI) developed one of the first frameworks to reduce inter-scanner differences for [^18^F]-fluorodeoxyglucose (FDG) imaging by post-reconstruction smoothing data to a pre-defined spatial resolution, as determined using a Hoffman phantom (*5*). There has been no definitive validation of this procedure for Aβ-PET, however. Further, we have demonstrated that post-reconstruction smoothing differentially affects Aβ- and Aβ+ groups (*6*). Since the ADNI initiative, several initiatives have recommended acquisition protocols and image quality procedures to standardize both Aβ-PET and FDG imaging for multicentre studies (*15,16*). Akamatsu *et al*. proposed a phantom procedure to optimise image quality of Aβ-PET and FDG imaging across different scanners, where participating sites choose reconstruction settings to meet predefined criteria for image noise, uniformity, contrast, and spatial resolution using phantom scans (*17*). A recent study proposed the use of an FDG-filled Hoffman phantom to define the upper and lower limits for recovery coefficients in grey matter and a grey-to-white matter ratio to standardise quantification across scanners, similar to the EARL initiative (*14*). The overhead associated with acquiring Hoffman phantom data and likelihood of experimental errors led Lodge *et. al*. to propose a simple method for calculating PET spatial resolution using the ^18^F-filled barrel phantom readily available as a part of routine quality assurance (*11*). These studies were based on phantom data; the impact of standardising quantification metrics has not to this point been evaluated on human scans.

In the current study, we hypothesised that multi-scanner Aβ-PET harmonisation can be achieved by matching spatial resolution as determined by the Lodge barrel phantom measurement. We interrogated this hypothesis using a three-camera comparison dataset of [^18^F]-NAV4694 radiotracer, where each participant was scanned on two out of a set of three scanners. Raw PET data and vendor-supplied PET image reconstruction toolboxes enabled comparison of a broad range of reconstruction settings and spatial resolutions. Degree of harmonisation was indicated by the similarity of participants’ Aβ-SUVR measures from each pair of scanners.

## METHODS

Three scanners were used in this study: i) Siemens Biograph Vision 600; ii) Siemens Biograph mCT; iii) Philips Gemini TF64. Hereinafter, these scanners are referred to as the Vision, mCT and Philips, respectively.

### Phantom spatial resolution calculation

A barrel phantom was used to measure the radial and axial PET spatial resolution of a given reconstruction configuration by calculating the full width at half maximum (FWHM) along the two directions, as proposed by Lodge *et. al*. (*11*): A barrel phantom filled with ^18^F was placed at the centre of the scanner field-of-view, with the long axis parallel to the axis of the scanner. One end of the phantom was lifted to create a small angle with the scanner axis. Phantom data was reconstructed with a set of reconstructions and uploaded to the Society of Nuclear Medicine and Molecular Imaging’s (SNMMI) phantom analysis toolkit (http://www.snmmi.org/PAT) to calculate the FWHM (average of axial and radial FWHMs) of each configuration. Hereinafter, ‘spatial resolution’ refers to the Lodge barrel phantom-computed spatial resolution.

### Data collection and image reconstructions

The use of three scanners allowed three pairwise comparisons between scanners: i) Vision-mCT; ii) mCT-Philips; iii) Vision-Philips. The same subjects were scanned for Aβ twice on the selected pair of scanners. All subjects were injected with [^18^F]-NAV4694 radiotracer 50 minutes prior to 20 minutes of continuous scanning. Ethics approval and consent to participate in this study were approved by the Austin Health Human Research Ethics Committee (HREC/18/Austin/201)

### Vision – mCT

Five subjects were scanned on both the Vision and mCT scanners with an average interscan interval of 45.6±4.27 weeks. Data were reconstructed with ordered subset expectation maximization with time-of-flight information (OPTOF). For the Vision, twelve iteration (*i*) values were used, {1, 2, 3, 4, 5, 6, 12, 18, 24, 30, 36, 42}, with the number of subsets, *s*=5. For the mCT, *s*=21 and *i* ε {2, 4, 6, 8, 10, 12}. No post-reconstruction Gaussian smoothing was used. The constant number of subsets reflected scanner software constraints. All scans were reconstructed at a voxel size of 1.65 mm × 1.65 mm × 2 mm.

### mCT – Philips

Eight subjects were scanned on both the mCT and Philips scanners with an inter-scan interval of 41.6±8.24 weeks. Philips data were reconstructed using one reconstruction configuration, the line-of-response row-action maximum likelihood (LOR RAMLA) with smoothing parameter set to ‘SHARP’ (the brain imaging protocol of the Philips T64). The spatial resolution of the Philips reconstruction configuration was 8.2 mm. mCT data were reconstructed using OPTOF with *i* ε {2, 4, 6, 8, 10, 12} and *s*=21. In order to further reduce the resolution of the mCT reconstructions, postreconstruction Gaussian smoothing was applied to the lowest resolution mCT reconstruction (*i*=2, *s*=21) from 1-13 mm in steps of 1 mm. All scans were reconstructed at a voxel size of 2 mm × 2 mm × 2 mm.

### Vision – Philips

Five subjects were scanned across both Vision and Philips scanners with an inter-scan interval of 31.66±6.98 weeks. Philips data were reconstructed using one reconstruction configuration, the line-of-response row-action maximum likelihood (LOR RAMLA) with smoothing parameter set to ‘SHARP’. Vision data were reconstructed using OPTOF with *i* ε{1, 2, 3, 4, 5, 6, 12, 18, 24, 30, 36, 42} and *s*=5. The lowest resolution achieved only changing the number of iterations was 9.1 mm. In order to further reduce the resolution of the Vision reconstructions, post-reconstruction Gaussian smoothing was applied to the lowest resolution Vision reconstruction (*i*=1, *s*=5) from 2-8 mm in steps of 2 mm. All scans were reconstructed at a voxel size of 2 mm × 2 mm × 2 mm.

### ADNI reconstruction comparison

ll data from the three scanners were reconstructed using ADNI-proposed reconstruction configurations: the mCT scanner data were reconstructed using 4*i*, 24*s* without enabling TOF information and post reconstruction smoothing; the Vision data were reconstructed without post-reconstruction smoothing using 8*i*,5*s* enabling TOF; Philips data were reconstructed using the Brain protocol with LOR-RAMLA, setting the smoothing parameter to ‘SHARP’ (*18*).

### Data Analysis

All scans were registered to a CT-based normalization template: The skull was stripped to remove off-target binding in non-brain regions. CT scans, acquired for attenuation correction, were used to generate a skull stripping mask using the FSL Brain Extraction Tool, and the mask was transformed to the PET domain. Results were manually checked for both registration and stripping faults. Each scan was nonlinearly registered to the FSL MNI152 1 mm brain template using Advanced Normalization Tools using the CT image as an intermediate step between the PET image and the template.

Aβ-PET SUVR values (hereinafter termed ‘SUVR’ values) were generated using the whole cerebellum defined by the standard CL method as the reference region (*19*). For each scan, SUVR values were computed across a total of 72 brain regions spread across the temporal lobe, frontal lobe, parietal lobe, cingulate and occipital lobe (Anatomical Labelling Atlas 3, AAL3) (*20*). For the Vision–mCT dataset, a total of 360 regional SUVR values (5 subjects × 72 brain regions) were extracted for each reconstruction configuration and compared. For the mCT–Philips dataset, the total was 576 regional SUVR values (8 subjects × 72 brain regions), and for the Vision – Philips dataset, 360 (5 subjects × 72 brain regions). Global SUVR values were also computed for each dataset.

### Assessment of harmonisation

Statistical similarity between SUVRs was assessed using paired (within subject) t-tests for each pair of reconstructions across scanners; the higher the p-value, the greater the similarity between the SUVRs. A second measure of harmonisation, that of root mean-squared-differences (RMSD) between paired global SUVR values, was employed.

For completeness, CapAIBL was used to generate CL values for all reconstructions on each set of data (http://milxcloud.csiro.au) (*21*).

## RESULTS

In order to test our hypothesis that harmonisation of Aβ-SUVR values between two scanners could be achieved by matching spatial resolutions, in the following we present SUVR similarity as a function of phantom-derived FWHM (resolution) and demonstrate that maximal SUVR harmonisation (statistical similarity) is achieved at matched spatial resolution. We further verify that matched spatial resolution provides SUVRs with minimal RMSD.

### Vision-mCT harmonization

The harmonization results between the mCT and Vision scanners are presented in **Figure 1**. The barrel phantom-derived spatial resolutions of the mCT reconstructions were between 4.30 mm and 4.85 mm. The Vision reconstruction with spatial resolution of 4.1 mm (6*i*, 5*s*, 0 mm smoothing) gave rise to maximal statistical similarity between regional SUVRs (**Figure 1. A**). The optimally harmonised reconstruction pair is marked in a black circle (**Figure 1. A, B**), and resulted in the minimal RMSD across all reconstruction configuration pairs (**Figure 1. B**). Paired global SUVRs calculated using the optimally harmonized reconstruction configuration pair were closely matched (**Figure 1. C)**.

**Figure 1:**
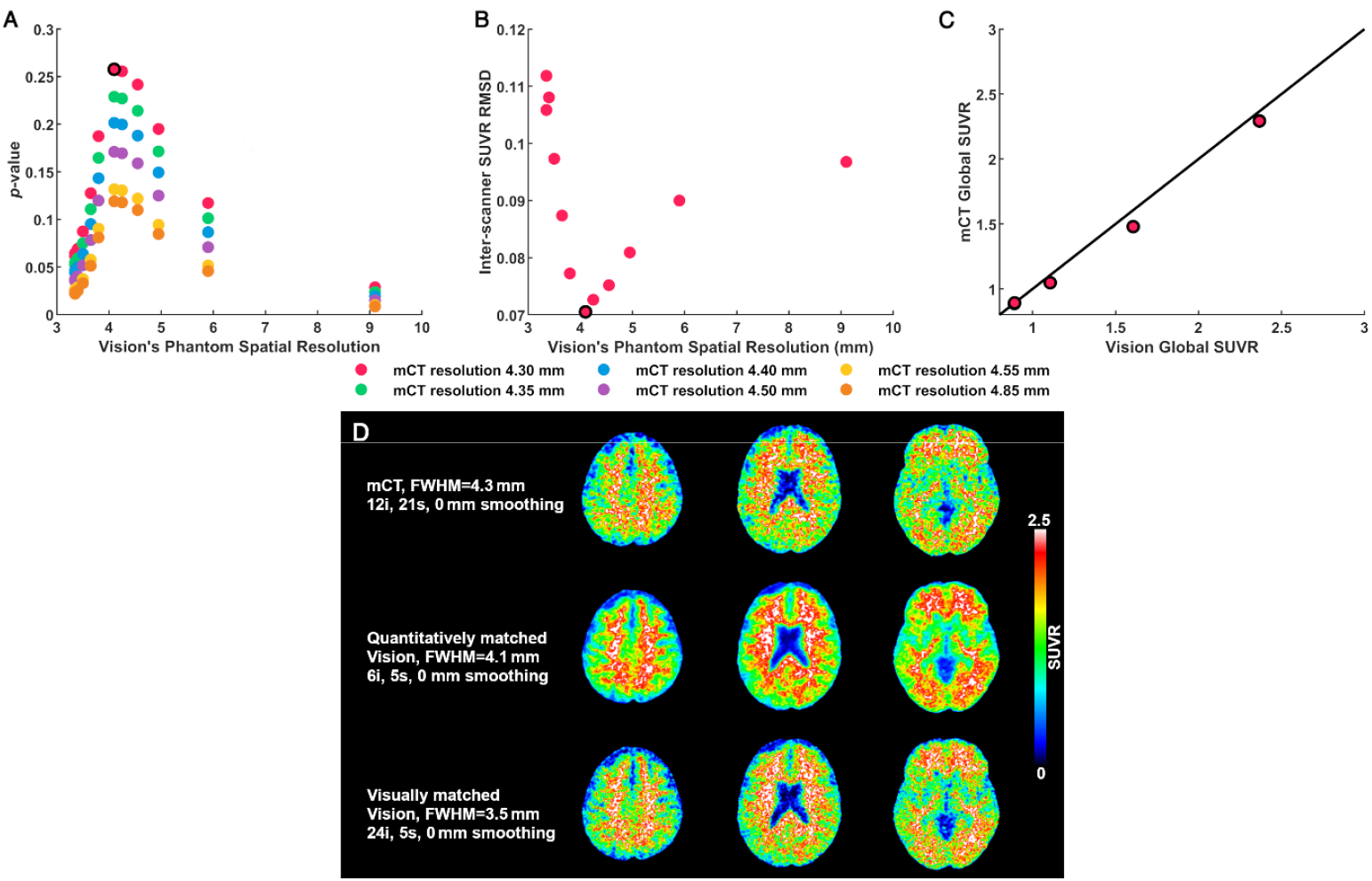
Vision-mCT harmonization. **A)** Statistical similarity between paired mCT-Vision regional SUVRs (higher *p*-value indicating greater similarity). **B)** Root mean square difference between paired mCT and Vision Global SUVRs (mCT: FWHM=4.3 mm across Vision FWHM resolutions). **C)** Global SUVR scatterplot for five subjects calculated using the harmonized reconstruction configuration pair. Four data points visible as two subjects’ data overlap at (0.89, 0.89). **D)** Visual comparison of the harmonization process between mCT and Vision using brain slices from a representative subject; First row: mCT reconstruction (FWHM=4.3 mm), second row: quantitatively-matched Vision reconstruction (FWHM=4.1 mm), third row: visually-matched Vision reconstruction (FWHM=3.5 mm). In panels **A)-C)**, black circles denote harmonised (maximally similar) pair of reconstruction configurations.

Representative brain slices from a subject scanned by both mCT and Vision are shown in **Figure 1. D**. The statistically matched mCT (FWHM=4.3 mm, 12*i*, 21*s*, 0 mm smoothing, 1^st^ row of **Figure 1. D**) and Vision (FWHM=4.1 mm, 24*i*, 5*s*, 0 mm smoothing, 2^nd^ row of **Figure 1. D**) reconstruction configuration are less visually similar than the Vision reconstruction configuration with FWHM=3.5 mm (24i, 5s, 0 mm smoothing, 3^rd^ row of **Figure 1. D**), demonstrating that visual matching between mCT and Vision does not quantitatively harmonize Aβ-SUVR values.

### mCT-Philips harmonization

**Figure** shows the harmonization results for the mCT-Philips scanner pair. There was only one standard brain mode clinical reconstruction algorithm for the Philips, with spatial resolution 8.2 mm. The mCT reconstruction configuration with FWHM=9.35 mm (2*i*, 21*s*, 8 mm smoothing) resulted in maximal similarity (highest *p*-value) between regional SUVRs (**Figure**. **A**). The RMSD between mCT and Philips global SUVRs across mCT resolutions demonstrates a broad trough with minimum greater than 9.35mm (**Figure**. **B)**. The eight subjects’ global SUVRs for the pair of harmonized reconstruction configuration were well matched (**Figure**. **C**).

Representative brain slices from a subject scanned by both mCT and Philips are depicted in **Figure**. **D**. The quantitatively harmonised Philips (FWHM=8.2 mm, 1^st^ row of **Figure**. **D**) and mCT (FWHM=9.35 mm, 2*i*, 21*s*, 8 mm smoothing, 2^nd^ row of **Figure**. **D**) reconstruction configurations are less visually similar than the mCT reconstruction configuration with FWHM=6.30 mm (2*i*, 21*s*, 4 mm smoothing, 3^rd^ row of **Figure**. **D**), again demonstrating the necessity of quantifiable metrics for harmonisation.

### Vision-Philips harmonization

Figure 2. shows the harmonization results for the Vision-Philips scanner pair. There was only one standard brain mode clinical reconstruction algorithm for the Philips, with spatial resolution 8.2 mm. The Vision reconstruction configuration with FWHM=9.1 mm (1*i*, 5*s*, 0 mm smoothing) resulted in maximal similarity (highest *p*-value) between regional SUVRs (**Figure 2. A**). The RMSD between Vision and Philips global SUVRs across Vision resolutions demonstrates a broad trough with minimum greater than 9.1mm (**Figure 2. B)**. The five subjects’ global SUVRs for the pair of harmonized reconstruction configurations were well matched (**Figure 2. C**).

**Figure 2:**
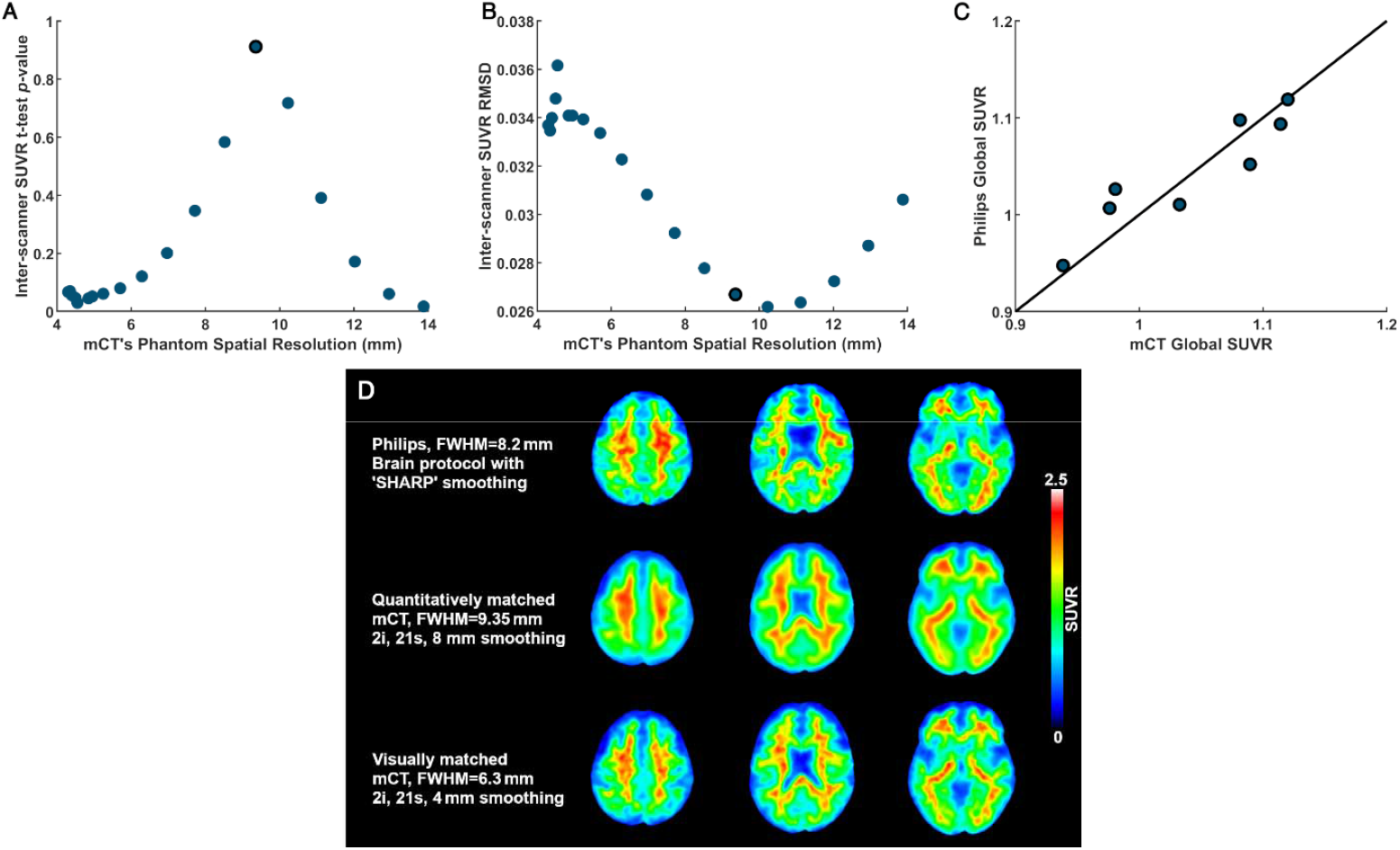
mCT-Philips harmonization. **A)** Statistical similarity between paired mCT-Philips regional SUVRs (higher *p*-value indicating greater similarity). **B)** Root mean square difference between paired mCT and Philips Global SUVRs (Philips: FWHM=8.2 mm across mCT FWHM resolutions). **C)** Global SUVR scatterplot for eight subjects calculated using the harmonized reconstruction configuration pair. **D)** Visual comparison of the harmonization process between mCT and Philips using brain slices from a representative subject; First row: Philips reconstruction (FWHM=8.2 mm), second row: quantitatively-matched mCT reconstruction (FWHM=9.35 mm), third row: visually-matched mCT reconstruction (FWHM=6.3 mm). In panels **A)-C)**, black circles denote harmonised (maximally similar) pair of reconstruction configurations.

**Figure 2:**
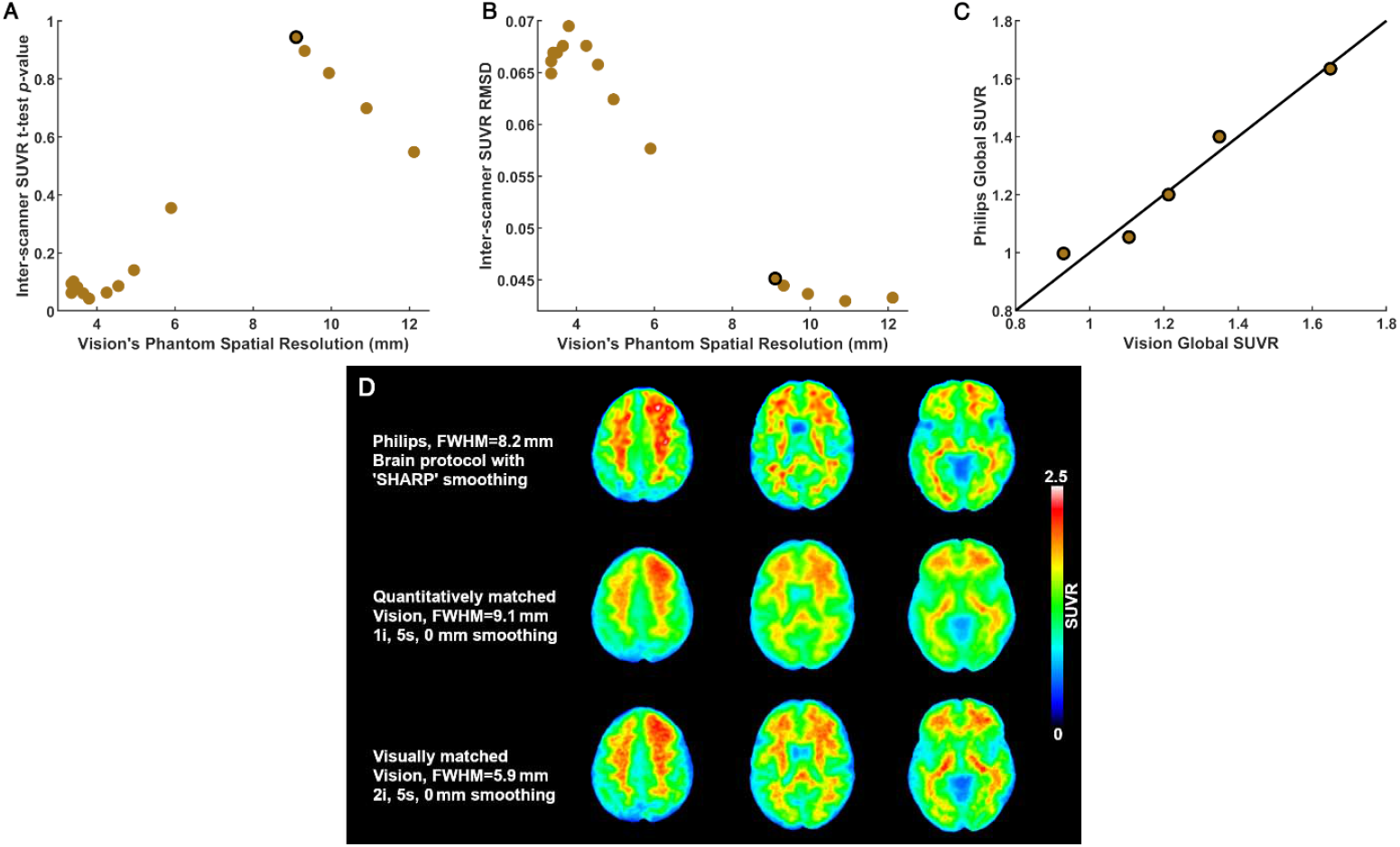
Vision-Philips harmonization. **A)** Statistical similarity between paired Vision-Philips regional SUVRs (higher *p*-value indicating greater similarity). **B)** Root mean square difference between paired Vision and Philips Global SUVRs (Philips: FWHM=8.2 mm across Vision FWHM resolutions). **C)** Global SUVR scatterplot for five subjects calculated using the harmonized reconstruction configuration pair. **D)** Visual comparison of the harmonization process between Vision and Philips using brain slices from a representative subject; First row: Philips reconstruction (FWHM=8.2 mm), second row: quantitatively-matched Vision reconstruction (FWHM=9.1 mm), third row: visually-matched Vision reconstruction (FWHM=5.9 mm). In panels **A)-C)**, black circles denote harmonised (maximally similar) pair of reconstruction configurations.

Representative brain slices from a subject scanned by both Vision and Philips are depicted in **Figure 2. D**. The quantitatively harmonised Philips (FWHM=8.2 mm, 1^st^ row of **Figure 2. D**) and Vision (FWHM=9.1mm, 1*i*, 5*s*, 0 mm smoothing, 2^nd^ row of **Figure 2. D**) reconstruction configurations are less visually similar than the Vision reconstruction configuration with FWHM=5.9 mm (2*i*, 5*s*, 0 mm smoothing, 3^rd^ row of **Figure 2. D**), again demonstrating the necessity of quantifiable metrics for harmonisation.

### Comparison with ADNI reconstructions

Regional SUVR-based statistical similarity and RMSD values of global SUVRs for ADNI reconstruction configurations are provided in **Supplementary Table S1**. The barrel phantom-derived FWHM values for ADNI reconstructions are 4.60 mm, 3.95 mm, and 8.2 mm for mCT, Vision and Philips, respectively. For the mCT-Vision scanner pair, regional SUVRs between the ADNI reconstructions were not significantly different (*p*=0.255) and RMSD between global SUVRs of 0.071. In contrast, regional SUVRs between the Philips and mCT reconstructions trended toward statistical significance (non-harmonised, *p*=0.058) and RMSD between global SUVRs of 0.031. Regional SUVR between the Vision and Philips were significantly different (non-harmonised, *p*=0.048) with RMSD between global SUVRs of 0.07.

### Centiloid harmonisation comparison

The CL scale, as a linear transformation of global SUVR values, should be harmonised for harmonised SUVRs. Comparisons of CL values calculated using the three sets of quantitatively harmonized scanner reconstruction pairs are given in **Supplementary Figure S1**. CL values between the mCT-Philips pair and between the Vision-Philips pair are well-matched. Comparison of mCT-Vision pair shows CL values that are larger for the Vision than mCT.

## DISCUSSION

In this study, we hypothesised that matching the spatial resolution of scanners, as measured by the Lodge barrel phantom method (*11*), harmonises Aβ-PET quantitation across scanners. This hypothesis was motivated by the knowledge that spatial resolution is affected by photon scattering, positron range of the radioisotope, hardware-specific limitations and choice of reconstruction algorithm and parameter configuration (*7,8*); consequently, partial volume effects affect PET quantitation (*6,9,10*). ADNI introduced the process of post-reconstruction smoothing to achieve the same spatial resolution across scanners to harmonize FDG brain scans (*5*). This procedure has not been validated for Aβ-PET imaging and our recent study has demonstrated that smoothing may not be optimal for matching the spatial resolution, as it changed the Aβ-PET SUVR in Aβ+ positive group and not in the Aβ-group (*6*).

Our results have provided significant evidence in support of the hypothesis of harmonisation via barrel phantom-derived spatial resolution, using two metrics of harmonisation: statistical similarity (*p*-value) between paired regional SUVRs and root mean square difference between paired global SUVRs. This is an important and promising outcome, as our proposed harmonisation procedure solely depends on phantom scans and no requirement for scanning subjects in multiple scanners. Further, the overhead associated with acquiring Hoffman phantom data is greatly reduced in the Lodge method based on an ^18^F-filled barrel phantom readily available in every centre as a part of routine quality assurance (*11*).

The two metrics of harmonisation, statistical similarity and RMSD, provided minor differences in harmonised reconstruction configurations for the Vision-Philips and mCT-Philips comparisons. This may be due to differences in the post-reconstruction smoothing processes employed by Philips and Siemens software; Siemens reconstructions used uniform smoothing throughout the brain while Philips used nonuniform region-based smoothing throughout the brain. Although our recent study suggests that post-reconstruction smoothing may not be optimal to harmonize Aβ-SUVR between scanners (*6*), in this research we applied smoothing in the harmonisation process between mCT and Philips scanners, out of necessity as the lowest resolution that could be achieved by the mCT by changing the number of iterations and subsets with TOF enabled was much higher than the Philips resolution. In future, it can be explored whether harmonization can be achieved without enabling the TOF in the reconstruction process, as it may provide a wider range of spatial resolution values. In the current study, TOF was enabled for every scanner to make reconstructions consistent.

For comparison, we implemented the ADNI-proposed reconstruction configurations for the mCT, Vision and Philips. They were shown to harmonize only the Aβ-PET SUVR quantitation between the mCT-Vision pair, and not the mCT-Philips and Vision-Philips pairs. This may be because ADNI reconstructions were proposed to standardise the PET image reconstruction across multiple centres; adhering to the proposed ADNI reconstruction configurations may effectively fulfil the intended purpose of standardising image reconstructions, however it may not achieve harmonization of Aβ-PET quantitation between the scanners.

We have further demonstrated that visual matching of Aβ-PET images across scanners does not robustly harmonize Aβ-PET quantitation. Our proposed process of matching spatial resolution has been shown to be more effective in harmonizing Aβ-PET quantitation. One possible explanation for this discrepancy is that visual matching focuses on the similarity of noise characteristics in the reconstructed images, which may not impact PVE between the white matter and grey matter of the brain. In contrast, spatial image resolution primarily governs PVE.

It is important to note that there is significant variability in the performance of PET scanners used in clinical settings, and the proposed harmonization scheme may compromise the high spatial resolution capabilities of newer systems. In our previous research, we observed that high-resolution imaging is beneficial in both cross-sectional and longitudinal Aβ-PET studies (*6*). As storing raw PET data for retrospective reconstruction can be a significant overhead in clinical settings, a potential solution is to reconstruct PET data in three different settings: high resolution for center-specific studies, a harmonized reconstruction configuration for multicenter-specific studies and a separate reconstruction for visual interpretation. This approach allows for the utilization of the most appropriate reconstruction method based on the context of the study.

Limitations of the current study include: (i) the mCT-Philips scanner comparison lacks Aβ+ positive subjects due to unavailability of participants; (ii) Only one Aβ-PET tracer was used, and the results may change with the level of off-target binding. Further studies can be explored in future to validate the proposed harmonization method on the other tracers; (iii) Analysis was done using one Aβ-PET processing pipeline; validation of several Aβ-PET pipelines could be conducted in future studies. Irrespective of these limitations, the results provide evidence in support of the hypothesis that barrel phantom-derived spatial resolution can be used for scanner harmonization in Aβ-PET imaging to reduce quantitation differences.

## CONCLUSION

Using Aβ-PET data from three sets of participants each scanned on a pair of PET scanners, we conclude that the process of matching spatial resolution measured by a barrel phantom can be used to minimize the Aβ-PET quantitation differences between the scanners. These promising results encourage application in Aβ-PET multi-centre trials and for PET camera updates during longitudinal studies.

## Data Availability

All data produced in the present study are available upon reasonable request to the authors

## ACKNOWLEDGMENTS

The authors acknowledge the facilities and scientific and technical assistance of the National Imaging Facility, a National Collaborative Research Infrastructure Strategy (NCRIS) capability, at the Melbourne Brain Centre Imaging Unit, the University of Melbourne. The first author would also like to acknowledge the Rowden White scholarship for its assistance in his research. The authors acknowledge the contributions of the the research teams of AIBL.

## KEY POINTS

### Question

Does the process of matching the barrel phantom-derived spatial resolution between scanners harmonize Aβ-SUVR quantitation?

### Pertinent findings

It has been validated that reconstruction pairs with matched barrel phantom-derived spatial resolution maximise the similarity between subjects’ paired Aβ-PET SUVR values recorded on two scanners.

### Implications for patient care

Harmonisation between scanners in multi-centre trials and PET camera updates in longitudinal studies can be achieved using a simple and efficient phantom measurement procedure, beneficial for the validity of Aβ-PET quantitation measurements.

